# Improving the detection of SARS-CoV-2 in point-of-care settings: a real-world performance evaluation of the Cobas SARS-CoV-2 nucleic acid test for use on the Cobas Liat System

**DOI:** 10.1101/2025.02.28.25323093

**Authors:** Myrto Bolanaki, Martin Möckel, Christopher Dodoo, Karen Gilliam, Elissa Robbins, D Jane Hata, Bruce White

**Affiliations:** Charité – Universitätsmedizin Berlin, Department of Emergency and Acute Medicine, Augustenburger Platz 1, 13353 Berlin, Germany; Roche Molecular Systems, Pleasanton, CA, USA; Department of Laboratory Medicine and Pathology, Mayo Clinic, Jacksonville, Florida, USA

**Keywords:** coronavirus disease 2019 (COVID-19), Cobas SARS-CoV-2 assay, Cobas Liat System, Cobas 6800/8800 System, point-of-care

## Abstract

**Background:** There is a need to confirm the clinical performance of point-of-care (POC) testing for severe acute respiratory coronavirus 2 (SARS-CoV-2) in emergency departments and time-sensitive outpatient settings. This study aimed to compare the clinical performance of the Cobas^®^ SARS-CoV-2 nucleic acid test for use on the Cobas Liat System (POC SARS-CoV-2) with the Cobas SARS-CoV-2 Qualitative Assay for use on the Cobas 6800/8800 System (68/8800) when used to detect SARS-CoV-2 infection in individuals at POC settings.

**Methods:** This prospective, two-site, non-interventional study, conducted in the US and Germany, collected fresh POC nasopharyngeal samples according to local procedures for upper respiratory swab sampling. Pairwise agreement was evaluated by estimating positive, negative, and overall percent agreement (PPA, NPA, and OPA, respectively) between POC SARS-CoV-2 and 68/8800 assays. Site-specific analyses were also conducted.

**Findings:** Overall, 317 evaluable samples were collected from March 30 to July 2023 (US site) and from November 7, 2023 to April 4, 2024 (German site). Relative to 68/8800, POC SARS-CoV-2 had a PPA of 98.8% (169/171) and an NPA of 90.4% (132/146). The OPA was 95.0%. Site-specific analyses were broadly comparable to the overall results, although enrichment for positive samples at the US site resulted in a PPA and NPA of 98.6% (139/141) and 55.6% (5/9), respectively.

**Interpretation:** POC SARS-CoV-2 used in real-world POC settings showed good overall clinical performance relative to 68/8800, a widely used and accurate laboratory-based test, for detecting SARS-CoV-2. Efficient POC testing will help improve the timely management of SARS-CoV-2 infections.

**Key points:**

- There are limited clinical studies evaluating the clinical performance of SARS-CoV-2 assays in real-world populations
- The Cobas^®^ SARS-CoV-2 nucleic acid test for use on the Cobas Liat System showed good clinical performance relative to a widely used laboratory-based test for detecting SARS-CoV-2 at two point-of-care hospital sites.
- Efficient point-of-care testing will help improve the timely management of SARS- CoV-2 infections

## 1. Introduction

Coronavirus disease 2019 (COVID-19), an acute respiratory illness caused by severe acute respiratory syndrome coronavirus-2 (SARS-CoV-2), was declared a global pandemic in March 2020 by the World Health Organization (WHO) following initial outbreaks in China and the subsequent spread to all regions of the globe [1–4]. The rapid spread of COVID-19 has resulted in >770 million cases of infection and >7 million deaths worldwide (as of 10 November 2024) [5], with the elderly and people with comorbidities being particularly affected [1,6].

Although the WHO no longer considers COVID-19 a global emergency, COVID-19 morbidity and mortality remain a serious concern [2,5,7]. Ongoing monitoring of COVID-19 remains key to improving its long-term management and mitigating its spread. Diagnosis of COVID-19 in people with suspected infection, particularly mild cases, cannot rely on typical clinical manifestations of respiratory infection alone (e.g., fever, cough, shortness of breath, myalgia, and fatigue) given the overlap of nonspecific signs and symptoms with other viral and bacterial conditions [8–10]. In cases of severe COVID-19, definitive diagnosis of SARS-CoV-2 infection should be sought in addition to laboratory and radiographic abnormalities to help healthcare providers (HCPs) confidently manage patients and the risk of local outbreaks [11]. Definitive diagnosis is often supported by real-time, reverse-transcription, polymerase chain reaction (RT-PCR) tests, a form of nucleic acid amplification technology (NAAT), to detect SARS-CoV-2 genetic material in respiratory specimens. These tests offer high accuracy in terms of clinical sensitivity and specificity in both symptomatic and asymptomatic patients [8,11,12]. High clinical sensitivity is key when screening asymptomatic patients for SARS-CoV-2 given the variability in viral load in this population [13,14]. Additionally, implementing RT-PCR testing at point of care (POC), instead of relying on central laboratories, allows for rapid turnaround times and detection of SARS-CoV-2 shortly after sample collection, when the likelihood of detectable viral loads may be at its highest [12,15]. POC testing plays a key role in SARS-CoV-2 management in emergency departments and time-sensitive outpatient settings, particularly when stat or on-demand testing is required, as well as in non-clinical settings, including long-term care facilities, retirement homes, schools, remote locations, and infection control screening points [8,12].

The Cobas^®^ SARS-CoV-2 nucleic acid test (Roche Diagnostics, USA) for use on the Cobas Liat System (herein referred to as POC SARS-CoV-2) employs RT-PCR technology to rapidly identify SARS-CoV-2 infection from nasopharyngeal or nasal swabs from individuals suspected of having respiratory viral infection, or from asymptomatic individuals, in both laboratory and POC settings with a turnaround time of approximately 20 minutes [16]. Given the urgent response required to address the COVID-19 pandemic, limited prospective clinical studies have been conducted to demonstrate the sensitivity and specificity of SARS-CoV-2 assays in real-world populations. Moreover, recent data suggest that molecular test results across platforms may be variable, reinforcing the need to confirm clinical performance [17,18]. The primary objective of this prospective, two-center study was to compare the clinical performance of POC SARS-CoV-2 with the Cobas SARS-CoV-2 Qualitative assay for use on the Cobas 6800/8800 System (Roche Diagnostics, USA; herein referred to as 68/8800) when used to detect SARS-CoV-2 infection in symptomatic and asymptomatic patients. The latter is a widely used high-throughput lab-based RT-PCR test [16] and serves as a robust reference method for the analysis.

## 2. Methods

### 2.1. Patient population

In this prospective, noninterventional study, fresh samples from asymptomatic or symptomatic patients with suspected SARS-CoV-2 infection were collected in a POC setting across two sites (Mayo Clinic, Jacksonville, Florida, USA and Charité University Hospital, Berlin, Germany). All SARS-CoV-2 tests were ordered by an HCP aligned with real-world standard-of-care (SOC) procedures. Suspicion of SARS-CoV-2 infection was defined as an individual with signs and/or symptoms consistent with respiratory infection (e.g., fever, cough, shortness of breath, or myalgia) or recent known exposure (i.e., close contact with an individual with confirmed COVID-19 or who had respiratory infection symptoms). Frozen, archived samples from individuals with suspected infection who had previously tested positive for SARS-CoV-2 were also used in the analysis.

### 2.2. Study ethics

The study was conducted in compliance with the International Council for Harmonization of Technical Requirements for Pharmaceuticals for Human Use (ICH) and Good Clinical Practice Guidelines. Institutional review board approval was obtained for each participating study site before study start (Ethics Committee of the Charité–Universitätsmedizin Berlin and Mayo Clinic Florida Institutional Review Committee). All participants provided written informed consent.

### 2.3. Sample collection and testing

During the study, the two participating sites used either the POC SARS-CoV-2 or 68/8800 tests as their standard method for detecting SARS-CoV-2. Clinical samples were prospectively collected at each site using their respective SOC procedures. Sample testing with POC SARS-CoV-2 was performed in accordance with the manufacturer’s assay instructions for use. All system operators were provided with the relevant user materials prior to the study and were blinded to all results from other test systems.

Eligible participants provided fresh nasopharyngeal samples. These were collected using each site’s established SOC procedures for upper respiratory swab sampling. Flocked swab samples were eluted in BD/Copan Universal Transport Medium (UTM), Remel Viral Transport Medium (VTM), or a similar 3-mL viral transport medium compatible with molecular testing for SARS-CoV-2. Testing of fresh, non-archived samples (3 mL sample; 200 µL for testing) was conducted on the Cobas Liat System as soon as possible, and up to a maximum of 4 hours, after sample collection. Subsequent 68/8800 testing was conducted at a central laboratory within 72 hours of collection (stored at 2–8°C).

The study protocol allowed for the use of archived specimens if prospectively collected samples were insufficient to reach the required number of positive samples. De-identified, frozen specimens were eligible for inclusion if they had previously tested positive (using any method) for SARS-CoV-2 and were collected in BD/Copan UTM, Remel VTM, or a similar viral transport media compatible with SARS-CoV-2 molecular testing (stored at −70°C or colder). A full list of specimen exclusion criteria is available in **Supplementary Table 1**.

Thawed samples were first retested for SARS-CoV-2 positivity if the previous test method for establishing SARS-CoV-2 positivity was not 68/8800. Following the 68/8800 test to establish SARS-CoV-2 positivity, POC SARS-CoV-2 was conducted within 4 hours for thawed samples stored at room temperature or within 4–24 hours for thawed samples stored at 2–8°C.

For both fresh and frozen specimens, repeat tests with either assay were conducted if an invalid result was generated in the first test; however, a second invalid result led to exclusion from the analysis.

### 2.4. Intended use of POC SARS-CoV-2 and 68/8800 assays

POC SARS-CoV-2 has been authorized for use under FDA Emergency Use Authorization (EUA), US 510(k) clearance and Conformité Européenne *in vitro* diagnostic (CE-IVD) in laboratory and POC settings for *in vitro* detection of SARS-CoV-2 in people with or without symptoms of COVID-19 [19,20]. In the US, it is also authorized for use at POC in settings operating under a Clinical Laboratory Improvement Amendments (CLIA) certificate of waiver, compliance, or accreditation [19]. Under FDA EUA and CE-IVD, the 68/8800 lab test is approved for *in vitro* detection of SARS-CoV-2 in individuals suspected of COVID-19 and those without symptoms [21–23].

### 2.5. Site-specific protocol variations

The real-world POC setting of this study gave rise to site-specific variation in some procedures not otherwise specified in the protocol; these were broadly related to sample handling and laboratory testing procedures that were instead conducted per site SOC. At the German site, all samples following POC SARS-CoV-2 testing were diluted for 68/8800 testing. Prior to dilution, a 600 µL aliquot was prepared and frozen (−20°C). Diluted samples consisted of ∼2.2 mL of the remaining original sample and 4.3 mL of added Cobas PCR Media. These diluted samples (stored at 2–8°C) were retested using POC SARS-CoV-2 before 68/8800 lab testing was completed. This was not the case at the US site where source (non-diluted) samples were used for both POC SARS-CoV-2 and 68/8800 testing as per manufacturer’s assay instructions for use.

At the US site, fresh specimens were initially processed using the Cobas SARS-CoV-2 & Influenza A/B test on the Cobas Liat System (Roche Diagnostics, USA); different than that stipulated by the study protocol. The same specimens were also processed on the Cobas 6800 System, which enabled the qualitative detection of SARS-CoV-2. Positive SARS-CoV-2 samples using this assay were archived according to SOC procedures. Following identification of the assay error, all positive samples were rerun following a single freeze– thaw cycle with POC SARS-CoV-2 as described above for frozen specimens (conducted at a different site: Mayo Clinic, Rochester, Minnesota, USA).

### 2.6. Outcomes and data analysis

The clinical performance of POC SARS-CoV-2 was evaluated relative to 68/8800. Subjects/specimens with both valid POC SARS-CoV-2 test results and valid 68/8800 test results were considered evaluable. Pairwise agreement between the two assays was evaluated by estimating the positive percent agreement (PPA), negative percent agreement (NPA), and overall percent agreement (OPA) with two-sided 95% confidence intervals (CI). PPA and NPA were calculated as the percentage of specimens with positive or negative results, respectively, on 68/8800 that also had positive or negative results on POC SARS-CoV-2.

Similarly, OPA was calculated as the percentage of total specimens with the same SARS-CoV-2 outcome in both assays. The distribution of cycle threshold (Ct) values for POC SAR- CoV-2 and 68/8800 were recorded for the exploratory analysis of discordant samples.

In terms of PPA, the acceptance criteria for POC SARS-CoV-2 were set at PPA ≥95% with a lower-bound 95% CI ≥88.5%. For NPA, the acceptance criteria were NPA ≥95% with a lower-bound 95% CI ≥90.0%. To achieve the defined PPA lower 95% CI boundary, an estimated total sample size of approximately 100 positive samples, out of a maximum of 1000 prospectively collected samples, was required for the analysis. A minimum of 60 positive samples (including fresh and frozen) was required for the analysis; there was no requirement for the minimum number of negative samples.

For the German site, clinical performance was also explored for the retested, diluted POC SARS-CoV-2 samples relative to 68/8800. In addition, pairwise agreement between the diluted POC SARS-CoV-2 samples and the corresponding source (non-diluted) samples was explored; statistical significance at p<0.05 was determined using a McNemar test.

Demographic characteristics, self-reported symptoms and vital signs were collected for each patient following enrolment or taken from electronic medical records; these data were summarized using descriptive statistics. Clinical raw study data were captured in Microsoft Excel, then transferred and analyzed using SAS/STAT^®^ software.

## 3. Results

### 3.1. Study population

A total of 317 samples were prospectively collected across the two sites (US site: n = 150; German site: n = 167). All samples were nasopharyngeal as determined by local procedures for each site. Sample collection at the US site occurred between March 30 and July 2023 (archived samples collected as early as January 26, 2023), while sample collection occurred between November 7, 2023 and April 4, 2024 at the German site. All samples were evaluable and included in the analysis. Of the 317 evaluable patients, 54.6% (n = 173) and 45.4% (n = 144) were assigned female and male at birth, respectively; 98.2% (n = 164) had reported symptoms with a mean (standard deviation [SD]) of 5.0 (4.5) days since symptom onset. The median (SD) age of the study population was 56.6 (19.3) years. Demographic characteristics at enrolment are summarized in **Table 1**. The most commonly self-reported symptoms at enrolment included cough (n = 79; 47.3%), fever or chills (n = 71; 42.5%), and difficulty breathing (n = 55; 32.9%) (**Supplementary Table 2**). Vital signs at enrolment were all within the normal clinical range (**Supplementary Table 3**).

**Table 1.** Demographics and baseline characteristics of evaluable subjects.

|  | **Evaluable subjects**<br>**(n = 317)** |
| --- | --- |
| **Age (years)** |  |
| Mean ± SD | 56.6 ± 19.30 |
| Median | 60.0 |
| Range | 4–92 |
| Unknown | 0 |
| **Age group (years), n (%)** |  |
| 0–18 | 6 (1.9%) |
| 19–54 | 120 (37.9%) |
| $\geq 55$ | 191 (60.3%) |
| **Sex, n (%)** |  |
| Male | 144 (45.4%) |
| Female | 173 (54.6%) |
| **SARS -CoV-2 vaccination status, n (%)** |  |
| Yes | 41 (24.6%) |
| No | 1 (0.6%) |
| Unknown | 125 (74.9%) |
| Not reported | 150 |
| **Has subject reported symptoms? n (%)** |  |
| Yes | 164 (98.2%) |
| No | 3 (1.8%) |
| Not reported | 150 |
| **Number of days since symptom onset?** |  |
| Mean ± SD | 5.0 ± 4.51 |
| Median | 3.0 |
| Range | 1–21 |
| Not reported | 201 |
| **Sample type, n (%)** |  |
| Nasopharyngeal | 317 (100.0%) |
| **Site, n (%)** |  |
| Germany | 167 (52.7%) |
| USA | 150 (47.3%) |

Subjects/specimens with both POC SARS-CoV-2 test results and valid 68/8800 test results were considered evaluable.
Unknown category indicates subjects for whom the corresponding information was not available.
SD, standard deviation.

### 3.2. Clinical performance – main analysis

For the overall clinical performance evaluation (**Table 2**), 167 fresh samples and 150 archived (frozen) samples were evaluable. These corresponded to the samples provided by the German and US sites, respectively. Of these 317 evaluable samples, 183 (prevalence: 57.7%, 95% Wilson score interval (WI) [52.1, 63.2]) and 171 (prevalence: 53.4%, 95% WI [0.48, 0.59]) tested positive for SARS-CoV-2 using POC SARS-CoV-2 and 68/8800, respectively. Two positive samples using 68/8000 were not detected by POC SARS-CoV-2. This resulted in a PPA (95% CI) of 98.8% (95.8, 99.7). Overall, 134 and 146 samples tested negative for SARS-CoV-2 using POC SARS-CoV-2 and 68/8800, respectively. Fourteen negative samples identified using 68/8800 were found positive by POC SARS-CoV-2. This resulted in an NPA (95% CI) of 90.4% (84.5, 94.2). The OPA (95% CI) was 95.0% (92.0, 96.9).

**Table 2.** Clinical performance comparison of the POC SARS-CoV-2 test with 68/8800 in all evaluable samples.

| POC SARS-CoV-2 test result | 68/8800 reference |  |  |
| --- | --- | --- | --- |
|  | Positive | Negative | Total |
| **Detected** | 169 | 14 | 183 |
| **Not detected** | 2 | 132 | 134 |
| **Total** | 171 | 146 | 317 |
| **PPA, % (95% CI)** | 98.8 (95.8, 99.7) |  |  |
| **NPA, % (95% CI)** | 90.4 (84.5, 94.2) |  |  |
| **OPA, % (95% CI)** | 95.0 (92.0, 96.9) |  |  |

CI, confidence interval; NPA, negative percent agreement; OPA, overall percent agreement; POC, point of care; PPA, positive percent agreement; SARS-CoV-2, severe acute respiratory syndrome coronavirus-2

**Table 3** provides an overview of the clinical performance of POC SARS-CoV-2 relative to 68/8800 by study site. At the US site, a PPA (95% CI) of 98.6% (95.0, 99.6) was observed in pairwise agreement analyses between POC SARS-CoV-2 and 68/8800. Out of 141 positive samples, 2 discordant false negative samples were recorded with POC SARS-CoV-2. Of note, out of the 150 samples collected at the US site and tested with 68/8800, the majority tested positive for SARS-CoV-2 (141/150 [94.0%]) leaving 9 negative samples available for the NPA analysis. Of these latter 9 negative samples, 4 discordant false positives were recorded with POC SARS-CoV-2. This yielded an NPA (95% CI) of 55.6% (26.7, 81.1). At the German site, the PPA (95% CI) was 100% (88.6, 100.0) with no discordance reported between assays, while NPA was 92.7% (87.1, 96.0) with 10 discordant false positives recorded with POC SARS-CoV-2 (**Table 3**). Of note, out of 167 samples collected at the German site and tested with 68/8800, 30 were positive (∼18%) while 137 were negative for SARS-CoV-2.

**Table 3.** Clinical performance comparison of the POC SARS-CoV-2 test with 68/8800 by study site.

| **Site** | **US site** | **German site** | **Overall** |
| --- | --- | --- | --- |
| **Total, N** | 150 | 167 | 317 |
| **TP / (TP+FN)** | 139 / 141 | 30 / 30 | 169 / 171 |
| **PPA, % (95% CI)** | 98.6<br>(95.0, 99.6) | 100<br>(88.6, 100.0) | 98.8<br>(95.8, 99.7) |
| **TN / (TN+FP)** | 5 / 9 | 127 / 137 | 132 / 146 |
| **NPA, % (95% CI)** | 55.6<br>(26.7, 81.1) | 92.7<br>(87.1, 96.0) | 90.4<br>(84.5, 94.2) |

CI, confidence interval; FN, false negative; FP, false positive; NPA, negative percent agreement; PPA, positive percent agreement; TN, true negative; TP, true positive.

### 3.3. Clinical performance – diluted samples

At the German site only, samples that were diluted (as per routine procedure) for 68/8800 testing were retested using POC SARS-CoV-2; this enabled an exploratory clinical performance analysis of 167 diluted evaluable samples (**Table 4**). Based on 30 positive SARS-CoV-2 samples identified using 68/8800, the PPA (95% CI) was 100.0% (88.6, 100.0) with no false negatives detected by POC SARS-CoV-2. Based on 137 negative SARS-CoV-2 samples identified using 68/8800, the NPA (95% CI) was 95.6% (90.8, 98.0) with 6 false positives detected by POC SARS-CoV-2. The OPA (95% CI) was 96.4% (92.4, 98.3). This repeat analysis of diluted samples was consistent with the pairwise agreement findings in the PPA main analysis; however, a slight increase in NPA from 92.7% to 95.6% was observed.

**Table 4.** Exploratory clinical performance comparison of the POC SARS-CoV-2 test with 68/8800 in diluted samples (German site only).

|  | **68/8800 reference** |  |  |
| --- | --- | --- | --- |
| **POC SARS-CoV-2 test result** | **Positive** | **Negative** | **Total** |
| **Detected** | 30 | 6 | 36 |
| **Not Detected** | 0 | 131 | 131 |
| **Total** | 30 | 137 | 167 |
| **PPA, % (95% CI)** | 100.0 (88.6,100.0) |  |  |
| **NPA, % (95% CI)** | 95.6 (90.8, 98.0) |  |  |
| **OPA, % (95% CI)** | 96.4 (92.4, 98.3) |  |  |

CI, confidence interval; NPA, negative percent agreement; PPA, positive percent agreement; OPA, overall percent agreement; SARS-CoV-2, severe acute respiratory syndrome coronavirus-2.

An exploratory pairwise agreement analysis between the diluted POC SARS-CoV-2 samples and the corresponding POC SARS-CoV-2 source (non-diluted) samples tested in the main analysis showed no statistically significant difference in terms of clinical performance between the two sets of samples (p = 0.157, McNemar test) (**Supplementary Table 4**).

### 3.4. Clinical performance – discordant samples

In the main analysis, the distribution profiles of Ct values for SARS-CoV-2 were broadly similar between POC SARS-CoV-2 and 68/8800 (median Ct [Ct range]: 19.9 [9.4–36.40] and 23.0 [15.3–36.1], respectively); however, the Ct distribution with POC SARS-CoV-2 appeared shifted to the left towards lower Ct values (**Figure 1**). Positive samples with Ct values <15 were only observed with POC SARS-CoV-2 (the lowest Ct value with 68/8800 was 15.5). The range of Ct values for the 14 false positives detected with POC SARS-CoV-2 was 31.5–36.4 (**Supplementary Table 5**), consistently near the mean limit of detection of the test (Ct 34.7). The two false negatives, although undetected with POC SARS-CoV-2, also had Ct values with 68/8800 near the limit of detection (36.1 and 34.5). Discordant samples from the German site accounted for ten false positive samples, while the US site accounted for four false positives and both false negatives (**Supplementary Table 5**).

**Figure 1.**
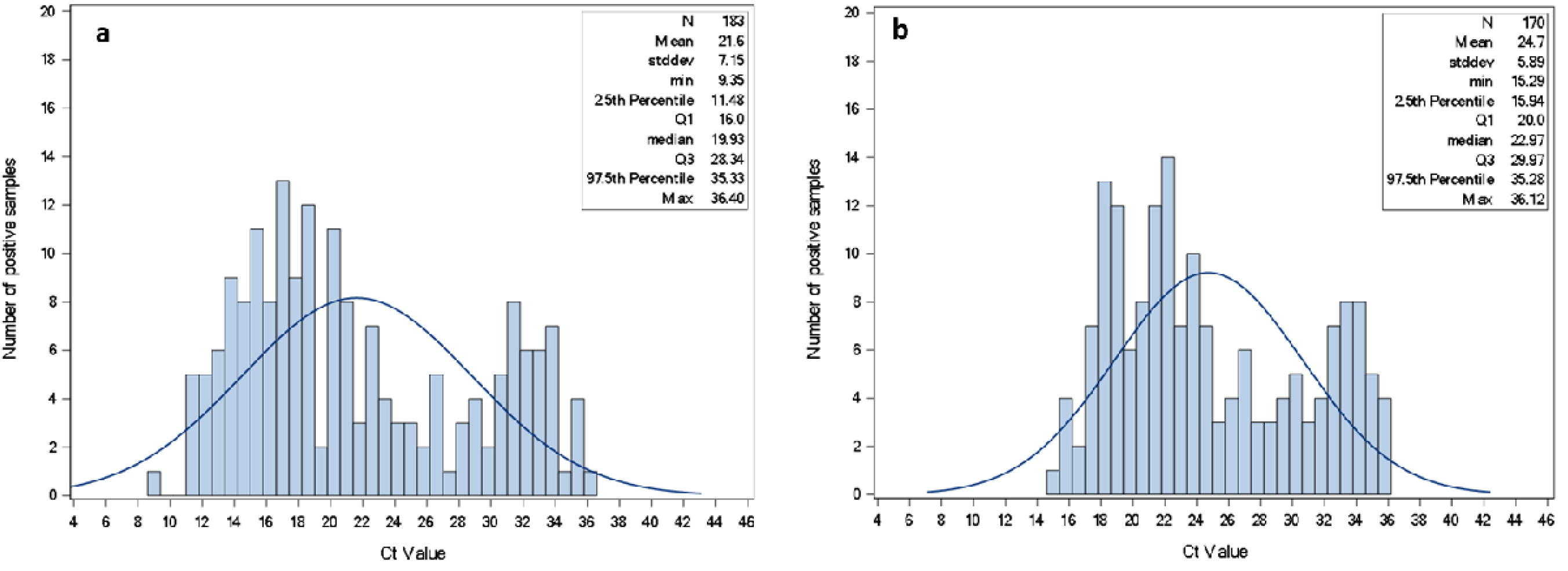
Distribution of Ct values for SARS-CoV-2 positive results with POC SARS-CoV-2 (a) and 68/8800 (b) Ct, cycle threshold; max, maximum; min, minimum; Q, quartile; stddev, standard deviation

The distribution profile of Ct values for the repeated POC SARS-CoV-2 test with diluted samples (German site only) is shown in **Supplementary** Figure 1. The Ct values for the six false positives identified in the diluted sample set ranged from 32.8 to 37.3. In this set of samples, four false positives were also identified in the main analysis; the remaining two false positives were SARS-CoV-2 negative in the main analysis with both assays.

## 4. Discussion

There is an unmet medical need for rapid, accurate, user-friendly, and near-patient or POC tests for SARS-CoV-2 detection, particularly in emergency care and time-sensitive outpatient settings [8,12]. Our two-center study evaluated the POC clinical performance of the POC SARS-CoV-2 assay to detect SARS-CoV-2 in individuals suspected of SARS-CoV-2 infection. Relative to the widely used 68/8800 test, a CE-IVD/FDA 510(k)-cleared lab-based NAAT, the POC SARS-CoV-2 assay achieved an overall PPA of 98.8%; this surpassed the predefined acceptance criteria threshold (≥95%) thus showing strong concordance between assays in terms of clinical sensitivity. An additional analysis of diluted samples (German site only) also showed strong pairwise alignment between assays with a PPA of 100%.

Conversely, the overall NPA was 90.4% with a lower 95% CI boundary of 84.5%; this did not meet predefined acceptance criteria thresholds (≥95% and lower-boundary of ≥90%) and was at odds with the expected NPA based on clinical studies that supported POC SARS-CoV-2 registration. In these studies, the clinical performance of POC SARS-CoV-2 against a composite comparator of three highly sensitive FDA EUA lab-based RT-PCR assays yielded an NPA (95% CI) of 99.5% (98.4, 99.8) and 99.6% (99.0, 99.8) in nasopharyngeal specimens from symptomatic and asymptomatic individuals, respectively [24].

The discrepant NPA result may be associated with the enrichment for SARS-CoV-2-positive specimens at the US site. This caused the overall number of negative samples to be much lower than anticipated based on SARS-CoV-2 positivity prevalence rates over the last year in the US and Europe (2–31%) [25,26]. Fresh specimens at the US site were initially processed using a different SARS-CoV-2 assay to that stipulated by the study protocol. Following this initial test, only those specimens that tested positive for SARS-CoV-2 were archived, resulting in an enriched archived sample of positive SARS-CoV-2 specimens. These archived specimens were then tested using POC SARS-CoV-2 according to the study protocol; however, of the 150 collected specimens, only 9 were SARS-CoV-2 negative (by 68/8800) and 4 of these 9 were discordant POC SARS-CoV-2 false positives. This yielded a site-specific, statistically underpowered NPA of 55.6%, which invariably drove the lower than anticipated overall NPA representative of both participating sites. The low number of negative samples at the US site may have also been driven by the source of sample collection i.e., individuals admitted to the emergency department who would have been overtly symptomatic of SARS-CoV-2 infection as the predominant respiratory virus circulating at the time. A more balanced sample of positive versus negative SARS-CoV-2 specimens would have increased the likelihood of achieving specificity values closer to the acceptance criteria. This is supported by the observed NPA values reported for the German site in the main analysis (92.7% [95% CI 87.1, 96.0]) and the improved NPA in the diluted analysis (95.6% [90.8, 98.0]) driven by a reduction in the number of false positives. Furthermore, a previous study using the Cobas Liat System, namely the Cobas SARS-CoV-2 & Influenza A/B assay, has also demonstrated PPA and NPA values >97% relative to 68/8800 in a similar study population [16].

To further investigate discordant results, Ct distribution profiles of SARS-CoV-2-positive specimens were explored. A modest leftward shift in the SARS-CoV-2 Ct distribution profile was observed with POC SARS-CoV-2 relative to 68/8800. In addition, the 14 false positives with POC SARS-CoV-2 in the main analysis (and the 6 false positives in diluted analysis) all had Ct values very close to the mean limit of detection (Ct 34.7). We cannot exclude the possibility that these samples were indeed SARS-CoV-2 positive but of very low viral load. Importantly, the reporting of discordant samples was similar between the two sites. In addition, the overall false positivity rate (4.4%) was close to the lower end of reported false positivity rates in other Cobas Liat System clinical performance studies (1.4–26.8% in symptomatic and asymptomatic individuals) [16,27,28]. These observations suggest that the POC SARS-CoV-2 assay may be slightly more sensitive than the 68/8800 assay, which became increasingly apparent when testing low viral load samples. This is consistent with a similar clinical performance study in which all discordant samples between the Cobas SARS-CoV-2 & Influenza A/B assay and 68/8800 were also near the limit of detection (range: 32.2– 37.4) [16]. We also cannot rule out the influence of other factors on the variability observed in Ct values and overall assay performance. All specimens in our study were nasopharyngeal, but subject age ranged from 4 to 92 years old (mean [SD]: 56.6 [19.30]) and number of days since symptom onset ranged from 1 to 21 days (mean [SD]: 5.0 [4.5]). Indeed, type of respiratory specimen; subject age; and quality, timing, and variability of sample collection have all been shown to influence the performance of POC tests [8].

A key limitation of this study was the inadvertent initial use of the multiplex Cobas SARS-CoV-2 & Influenza A/B assay at the US site. This led to a different set of specimen conditions between the two study sites (e.g., transport, storage, time of analysis), in addition to the aforementioned enrichment for SARS-CoV-2 positivity, when testing was conducted with POC SARS-CoV-2. Nevertheless, PPA was confirmed at both sites and the number of discordant samples were similar between sites. Although NPA could not be confirmed because of the overall low number of negative samples (and therefore statistically underpowered analysis), the overall capability of POC SARS-CoV-2 to detect SARS-CoV-2 in a real-world setting remains important. Moreover, the benefits of the POC SARS-CoV-2 assay (e.g., small footprint, easy-to-use interface, automation, rapid detection) enabled its rapid deployment in a POC setting, which is key when striving to achieve the ideal diagnostic assay as defined by WHO ASSURED criteria (Affordable, Sensitive, Specific, User-friendly, Rapid/Robust, Equipment-free, Deliverable) [15,29,30]. Our study was conducted at two geographically diverse sites (US and Germany) and reflect real-world SARS-CoV-2 testing practice; however, additional real-world analyses are warranted in developing countries to evaluate the performance of the assay in hands of local operators relative to local routine lab tests. Further analyses in asymptomatic individuals are also warranted given that only three asymptomatic individuals participated in our study; this, however, is indeed a reflection of real-world circumstances. An additional limitation may be the absence of self-collected nasal swabs during our study. Although the protocol allowed for both nasal and nasopharyngeal swabs, only HCP-collected nasopharyngeal specimens were collected in practice.

Nevertheless, clinical studies that supported POC SARS-CoV-2 registration demonstrated high clinical performance in nasal swab specimens comparable to that of nasopharyngeal specimens [24]. Analyses beyond clinical performance were not possible as individual patient-level data were not collected during the study.

Although COVID-19 may no longer be considered a global emergency, the need to rapidly detect SARS-CoV-2 and monitor transmission levels remains of critical importance, particularly in places with limited healthcare infrastructure. Over 250 SARS-CoV-2 NAAT tests have received EUA by the FDA [31]; however, robust performance studies in real-world populations to support these tests are lacking. In our study, although additional real-world analyses are needed to further validate the clinical specificity of POC SARS-CoV-2, we showed that POC SARS-CoV-2 used in real-world POC settings has good overall clinical performance in relation to the 68/8800 reference, a widely used and accurate lab-based RT-PCR test, with an OPA (95% CI) of 95.0% (92.0, 96.9) for detecting SARS-CoV-2.

## Data Availability

The study was conducted in accordance with applicable regulations. There are ethical and legal restrictions on sharing the de-identified data set used for our analysis. Any access requests from qualified researchers should be submitted directly to the Ethical Committee of each participating study site.

## Acknowledgments

The authors would like to express their gratitude to all participants for their valuable contributions to this study. Special thanks are extended to all personnel involved, particularly te dedicated study nurses of the Department of Emergency Medicine at Campus Virchow-Klinikum, as well as laboratory technician Fabian Holert. We would also like to acknowledge the staff of the Mayo Clinic Florida Clinical Microbiology Laboratory, and Jessica Tufariello, Associate Project Manager at the Mayo Clinic Rochester site. Third-party medical writing assistance, under the direction of the authors, was provided by Steven Barberini, PhD, The Salve Health Ltd, UK, and was funded by Roche Molecular Systems.

## Statements and Declarations

### Data availability statement

The study was conducted in accordance with applicable regulations. There are ethical andlegal restrictions on sharing the de-identified data set used for our analysis. Any access requests from qualified researchers should be submitted directly to the Ethical Committee of each participating study site.

### Author contributions

All authors contributed to the study conception and design. Data collection was performed by MB, MM, JH, and BW. Data analysis was conducted by CD. All authors were involved in drafting the manuscript. All authors read and approved the final manuscript.

## References

1. Chen N, Zhou M, Dong X, Qu J, Gong F, Han Y. Epidemiological and clinical characteristics of 99 cases of 2019 novel coronavirus pneumonia in Wuhan, China: a descriptive study. Lancet. 2020;395(10223):507–13. 10.1016/S0140-6736(20)30211-7

2. World Health Organization. Coronavirus disease (COVID-19) pandemic. https://www.who.int/europe/emergencies/situations/covid-19. Accessed 27 Nov 2024.

3. Wolfel R, Corman VM, Guggemos W, Seilmaier M, Zange S, Müller MA, et al. Virological assessment of hospitalized patients with COVID-2019. Nature. 2020;581(7809):465–9. 10.1038/s41586-020-2196-x

4. Zhu N, Zhang D, Wang W, Li X, Yang B, Song J,et al. A novel coronavirus from patients with pneumonia in China, 2019. N Engl J Med. 2020;382(8):727–33. 10.1056/NEJMoa2001017

5. World Health Organization. WHO Coronavirus (COVID-19) Dashboard. https://covid19.who.int/. Updated 10 Nov 2024. Accessed 27 Nov 2024.

6. Guan WJ, Ni ZY, Hu Y, Liang W-H, Ou C-Q, He J-X, et al. Clinical characteristics of coronavirus disease 2019 in China. N Engl J Med. 2020;382(18):1708–20. 10.1056/NEJMoa2002032

7. Bajema KL, Bui DP, Yan L, Li Y, Rajeevan N, Vergun R, et al. Severity and long-term mortality of COVID-19, influenza, and respiratory syncytial virus. JAMA Intern Med. 2025; 10.1001/jamainternmed.2024.7452

8. Basile K, Kok J, Dwyer DE. Point-of-care diagnostics for respiratory viral infections. Expert Rev Mol Diagn. 2018;18(1):75–83. 10.1080/14737159.2018.1419065

9. Ding Q, Lu P, Fan Y, Xia Y, Liu M. The clinical characteristics of pneumonia patients coinfected with 2019 novel coronavirus and influenza virus in Wuhan, China. J Med Virol. 2020;92(9):1549–55. 10.1002/jmv.25781

10. Liang WH, Guan WJ, Li CC, Li Y, Liang H, Zhao Y, et al. Clinical characteristics and outcomes of hospitalised patients with COVID-19 treated in Hubei (epicentre) and outside Hubei (non-epicentre): a nationwide analysis of China. Eur Respir J. 2020;55(6):2000562. 10.1183/13993003.00562-2020

11. Abbasi-Oshaghi E, Mirzaei F, Farahani F, Khodadadi I, Tayebinia H. Diagnosis and treatment of coronavirus disease 2019 (COVID-19): Laboratory, PCR, and chest CT imaging findings. Int J Surg. 2020;79:143–53. 10.1016/j.ijsu.2020.05.018

12. Caliendo AM, Gilbert DN, Ginocchio CC, Hanson KE, May L, Quinn TC, et al. Better tests, better care: improved diagnostics for infectious diseases. Clin Infect Dis. 2013;57(Suppl 3):S139–70. 10.1093/cid/cit578

13. Mozgovoj M, Graham M, Ferrufino C, Blanc S, Souto AF, Pilloff M, et al. Viral load in symptomatic and asymptomatic patients infected with SARS-CoV-2. What have we learned? J Clin Virol Plus. 2023;3(4):100166. 10.1016/j.jcvp.2023.100166

14. Viswanathan M, Kahwati L, Jahn B, Giger K, Dobrescu AI, Hill C, et al. Universal screening for SARS-CoV-2 infection: a rapid review. Cochrane Database Syst Rev. 2020;9(9):Cd013718. 10.1002/14651858.CD013718

15. McPartlin DA, O’Kennedy RJ. Point-of-care diagnostics, a major opportunity for change in traditional diagnostic approaches: potential and limitations. Expert Rev Mol Diagn. 2014;14(8):979–98. 10.1586/14737159.2014.960516

16. Hansen G, Marino J, Wang ZX, Beavis KG, Rodrigo J, Labog K, et al. Clinical performance of the point-of-care cobas Liat for detection of SARS-CoV-2 in 20 minutes: a multicenter study. J Clin Microbiol. 2021;59(2):e02811–20. 10.1128/jcm.02811-20

17. Fukasawa LO, Sacchi CT, Gonçalves MG, Lemos APS, Almeida SCG, Caterino-de-Araujo A. Comparative performances of seven quantitative reverse-transcription polymerase chain reaction assays (RT-qPCR) for detecting SARS-CoV-2 infection in samples from individuals suspected of COVID-19 in São Paulo, Brazil. J Clin Virol Plus. 2021;1(1):100012. 10.1016/j.jcvp.2021.100012

18. Munir R, Scott LE, Noble LD, Steegen K, Hans L, Stevens WS. Performance evaluation of four qualitative RT-PCR assays for the detection of severe acute respiratory syndrome coronavirus 2 (SARS-CoV-2). Microbiol Spectr. 2023;11(2):e03716–22. 10.1128/spectrum.03716-22

19. Roche Diagnostics. cobas® SARS-CoV-2. Nucleic acid test for use on the cobas® liat system. https://diagnostics.roche.com/global/en/products/lab/cobas-sars-cov-2-liat-rmd-liat-sars-003.html#productInfo. Updated 12 Dec 2024. Accessed 12 Dec 2024.

20. US Food and Drug Administration. 510(k) Substantial equivalence determination decision summary. cobas SARS-CoV-2 nucleic acid test for use on the cobas Liat System. https://www.accessdata.fda.gov/cdrh_docs/reviews/K223783.pdf. Accessed 16 Jan 2024.

21. Roche Diagnostics. cobas SARS-COV-2. Qualitative assay for use on the cobas 6800/8800 Systems. Instructions for use. https://www.fda.gov/media/136049/download?attachment. Updated 30 Aug 2024. Accessed 12 Dec 2024.

22. US Food & Drug Administration. Emergency Use Authorization. cobas® SARS-CoV-2. Qualitative assay for use on the cobas® 6800/8800 Systems. 2021. https://www.fda.gov/media/136046/download?attachment. Accessed 26 Feb 2025.

23. Roche Diagnostics. CE IVD. cobas® SARS-CoV-2 Qualitative. Nucleic acid test for use on the cobas® 6800/8800 Systems. 2021. https://elabdoc-prod.roche.com/eLD/api/downloads/c0618754-836d-ec11-0f91-005056a71a5d?countryIsoCode=XG. Accessed 26 Feb 2025.

24. Roche Diagnostics. cobas SARS-CoV-2 Nucleic acid test for use on the cobas Liat System - English - US-IVD. https://elabdoc-prod.roche.com/eLD/api/downloads/fe6bf2b6-dd87-ef11-2691-005056a772fd?countryIsoCode=XG. Updated 10 Nov 2024. Accessed 12 Dec 2024.

25. Centre for Disease Control and Prevention. COVID Data Tracker. Trends in United States COVID-19 deaths, emergency department (ED) visits, and test positivity by geographic area. https://covid.cdc.gov/covid-data-tracker/#trends_weeklydeaths_testpositivity_00. Updated 9 Dec 2024. Accessed 12 Dec 2024.

26. European Centre for Disease Prevention and Control. Overview of respiratory virus epidemiology in the EU/EEA. https://erviss.org/. Updated 1 Dec 2024. Accessed 12 Dec 2024.

27. Blackall D, Moreno R, Jin J, Plotinsky R, Dworkin R, Oethinger M. Performance characteristics of the Roche Diagnostics cobas Liat PCR System as a COVID-19 screening tool for hospital admissions in a regional health care delivery system. J Clin Microbiol. 2021;59(10):e0127821. 10.1128/jcm.01278-21

28. Egerer R, Edel B, Hornung F, Deinhardt-Emmer S, Baier M, Lewejohann J-C, et al. SARS-CoV-2 testing of emergency department patients using cobas^®^ Liat^®^ and eazyplex^®^ rapid molecular assays. Diagnostics (Basel). 2023;13(13):2245. 10.3390/diagnostics13132245

29. Naseri M, Ziora Z, Simon G, Batchelor W. ASSURED-compliant point-of-care diagnostics for the detection of human viral infections. Rev Med Virol. 2021;32(2):e2263. 10.1002/rmv.2263

30. Land KJ, Boeras DI, Chen XS, Ramsay AR, Peeling RW. REASSURED diagnostics to inform disease control strategies, strengthen health systems and improve patient outcomes. Nat Microbiol. 2019;4(1):46–54. 10.1038/s41564-018-0295-3

31. US Food & Drug Administration. In vitro diagnostics EUAs - Molecular diagnostic tests for SARS-CoV-2. 2023. https://www.fda.gov/medical-devices/covid-19-emergency-use-authorizations-medical-devices/in-vitro-diagnostics-euas-molecular-diagnostic-tests-sars-cov-2. Updated 19 Nov 2024. Accessed 27 Nov 2024.

