## Supplementary Appendix for "Improving the detection of SARS-CoV-2 in point-of-care settings: a real-world performance evaluation of the Cobas SARS-CoV-2 nucleic acid test for use on the Cobas Liat System"

**Affiliations:**

**Supplementary Table 1.** Sample exclusion criteria

- Sample volumes insufficient for testing:
    - For fresh non-archived samples, remaining volume after initial POC SARS-CoV-2 testing <2 mL
    - For frozen archived samples, post-thawing remnant volume <1.5 mL
- Other sample types (e.g., saliva, oropharyngeal, nasal/nasopharyngeal wash, sputum, bronchoalveolar lavage)
- Collection into other transport media types (e.g., Liquid Amies, saline)
- Storage outside the predefined stability requirements

**Supplementary Table 2.** Self-reported symptoms in evaluable patients.

| Symptoms | Result category (yes/no) | n (%) |
|---|---|---|
| **Fever or chills** | Yes | 71 (42.5) |
| | No | 96 (57.5) |
| **Shortness of breath** | Yes | 48 (28.7) |
| | No | 119 (71.3) |
| **Difficulty breathing** | Yes | 55 (32.9) |
| | No | 112 (67.1) |
| **New loss of taste or smell** | Yes | 0 (0) |
| | No | 167 (100.0) |
| **Congestion or runny nose** | Yes | 14 (8.5) |
| | No | 151 (91.5) |
| **Diarrhea** | Yes | 23 (13.9) |
| | No | 142 (86.1) |
| **Cough** | Yes | 79 (47.3) |
| | No | 88 (52.7) |
| **Fatigue** | Yes | 45 (27.1) |
| | No | 121 (72.9) |
| **Headache** | Yes | 31 (18.6) |
| | No | 136 (81.4) |
| **Sore throat** | Yes | 27 (16.2) |
| | No | 140 (83.8) |
| **Nausea or vomiting** | Yes | 36 (21.6) |
| | No | 131 (78.4) |
| **Muscle or body aches** | Yes | 31 (18.6) |
| | No | 136 (81.4) |

**Supplementary Table 3.** Vital signs of enrolled participants.

| Symptoms | Parameter | Total |
|---|---|---|
| **Heart rate, beats per minute** | N | 155 |
| | Mean ± SD | 92.1 ± 19.84 |
| | Median | 89.0 |
| | Range | 46.0–150.0 |
| | Not available | 162 |
| **Respiratory rate, breaths per minute** | N | 112 |
| | Mean ± SD | 18.0 ± 4.30 |
| | Median | 16.0 |
| | Range | 12.0–33.0 |
| | Not available | 205 |
| **Pulse oximetry ($SpO_2$), %** | N | 158 |
| | Mean ± SD | 97.2 ± 3.29 |
| | Median | 98.0 |
| | Range | 79.0–100.0 |
| | Not available | 159 |
| **Temperature, °C** | N | 148 |
| | Mean ± SD | 37.1 ± 0.94 |
| | Median | 37.0 |
| | Range | 34.7–39.9 |
| | Not available | 169 |

SD, standard deviation; $SpO_2$, oxygen saturation.

**Supplementary Table 4**. Exploratory clinical performance comparison of POC SARS-CoV-2 diluted samples with POC SARS-CoV-2 source (non-diluted) samples.

| | **POC SARS-CoV-2 Test Result - Undiluted** | | | |
|---|---|---|---|---|
| **POC SARS-CoV-2 Test Result- Diluted** | **Detected** | **Undetected** | **Total** | **P value** |
| **Detected** | 34 | 2 | 36 | 0.157 |
| **Not Detected** | 6 | 125 | 131 | |
| **Total** | 40 | 127 | 167 | |
| **PPA (95% CI)** | 85.0% (70.9%, 92.9%) | | | |
| **NPA (95% CI)** | 98.4% (94.4%, 99.6%) | | | |
| **OPA (95% CI)** | 95.2% (90.8%, 97.6%) | | | |

CI, confidence interval; NPA, negative percent agreement; OPA, overall percent agreement; POC, point of care; PPA, positive percent agreement; SARS-CoV-2, severe acute respiratory syndrome coronavirus-2.

**Supplementary Table 5. Ct values for POC SARS-CoV-2 and 68/8800 discordant samples.**

| Site | POC SARS-CoV-2 | | 68/8800 | |
|---|---|---|---|---|
| | Test result | Ct value | Test result | Ct value |
| *Main analysis* | | | | |
| US | Detected | 35.4 | Not detected | |
| US | Detected | 33.5 | Not detected | |
| US | Detected | 33.01 | Not detected | |
| US | Detected | 34.52 | Not detected | |
| US | Not detected | | Detected | 36.12 |
| US | Not detected | | Detected | 34.53 |
| Germany | Detected | 35.57 | Not detected | |
| Germany | Detected | 32.07 | Not detected | |
| Germany | Detected | 36.4 | Not detected | |
| Germany | Detected | 34.11 | Not detected | |
| Germany | Detected | 33.97 | Not detected | |
| Germany | Detected | 31.54 | Not detected | |
| Germany | Detected | 33.64 | Not detected | |
| Germany | Detected | 35.74 | Not detected | |
| Germany | Detected | 33.38 | Not detected | |
| Germany | Detected | 33.45 | Not detected | |
| *Dilution analysis (German site only)* | | | | |
| Germany | Detected | 34.11 | Not detected | |
| Germany | Detected | 35.1 | Not detected | |
| Germany | Detected | 32.84 | Not detected | |
| Germany | Detected | 35.32 | Not detected | |
| Germany | Detected | 37.25 | Not detected | |
| Germany | Detected | 33.35 | Not detected | |

Ct, cycle threshold; POC, point of care; SARS-CoV-2, severe acute respiratory syndrome coronavirus-2.

**Supplementary Figure 1.** Distribution of Ct values for SARS-CoV-2 positive results for diluted samples on POC SARS-CoV-2.

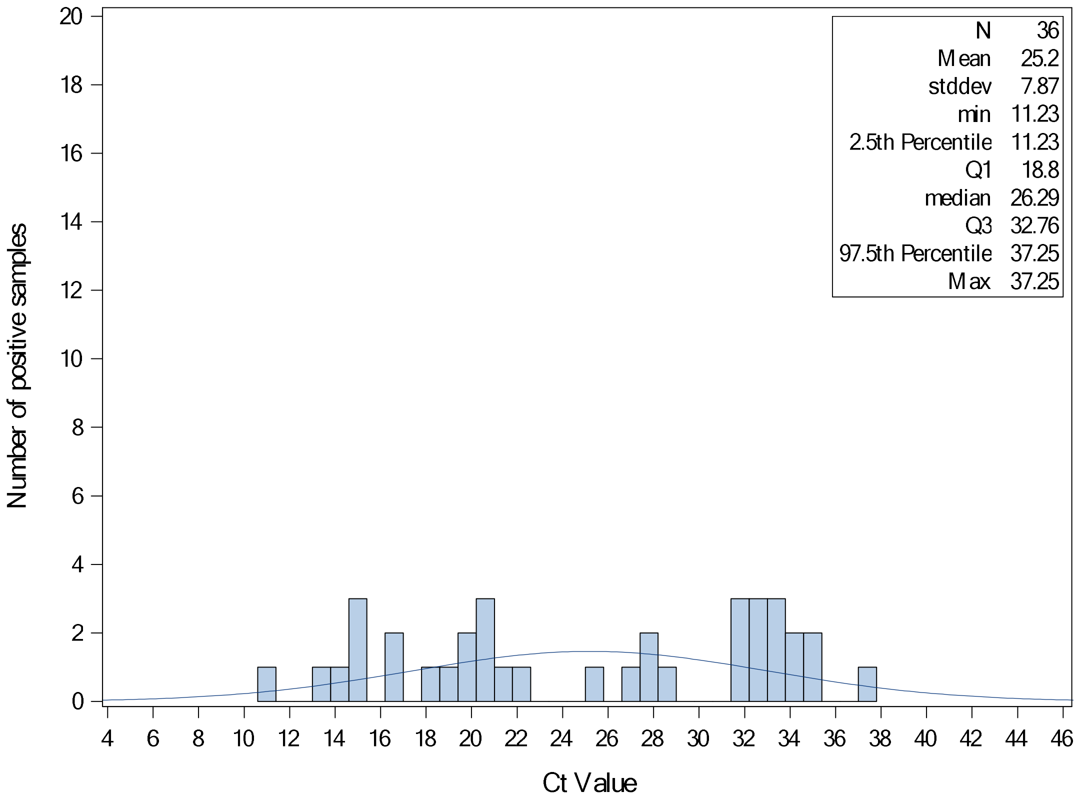
